# Alterations in the Brain Lipidome of Alzheimer’s Disease Donors with Rare TREM2 Risk Variants

**DOI:** 10.1101/2024.08.22.24311238

**Authors:** Petroula Proitsi, Amera Ebshiana, Asger Wretlind, Jin Xu, Angela Hodges, Cristina Legido-Quigley

## Abstract

TREM2 is a microglial receptor, sensitive to Phospholipids and Sphingomyelins, associated with neurodegeneration. Hypomorphic variants in the *TREM2* gene significantly increase the risk of developing AD.

The main aim of this study was to characterize networks of lipids in post-mortem brain from AD and control donors, and to identify key lipids that are associated with AD and impacted by dysfunctional TREM2.

We studied human post-mortem brain tissue from the hippocampus and the BA9 pre-association cortex from a total of 102 post-mortem brains. Specifically, brain tissue from the BA9 pre-association cortex was available for n=55 donors and brain tissue from the hippocampus was available for n=47 brain donors from three groups: AD donors with a non-synonymous risk DNA variant in *TREM2* (AD-TREM2^var^), AD donors with no *TREM2*-associated variant (AD-TREM2^wt^), and control donors. Mass Spectrometry was performed to obtain lipidomic signatures spanning 99 lipid species that included the following lipid classes: Ceramides, Sphingomyelins, Phosphatidic acids, Phopshatidyl-cholines, Phosphatidyl-ethalonamines, Phosphatidyl-glycerols, Phosphatidyl-inositols, Phosphatidyl-serines and Triglycerides. Weighted gene co-expression network analysis (WGCNA) was used to identify highly correlated lipid modules and ‘hub’ lipids. Linear mixed models and linear regression analyses, adjusted for age, biological sex, number of APOEε4 alleles and post-mortem delay were used to assess the association of modules and individual hub lipids with AD and *TREM2* status.

Four lipid modules were found to be relatively well-preserved between the two brain regions, and three of these modules were altered in AD donors and/or in AD donors with *TREM2* genetic variants. Levels of the BA9 “turquoise” module (“blue” hippocampus module), enriched in Sphingolipids and Phospholipids, were elevated in AD brain donors and in AD TREM2 carriers. The key lipid (hub) of the BA9 “turquoise”/ hippocampus “blue” module was a Phosphatidylserine (PS(32:1)), increased in both AD donors and TREM2 carriers (AD-TREM2^wt^ versus controls: beta=0.468, 95% CI 0.05 – 0.89, p= 3.02E-02; and AD-TREM2^var^ versus controls: beta=1.00, 95% CI 0.53 – 1.47, p= 5.57E-03), whereas the strongest association was observed with a Ceramide (Cer(d38:1)) (AD-TREM2^wt^ versus controls: beta=0.663, 95% CI 0.17 – 1.16, p= 8.9E-03; and AD-TREM2^var^ versus controls: beta=1.31, 95% CI 0.78 – 1.84, p= 4.35E-06).

The consistent increase in TREM2 ligands such as Ceramides and Phosphatidyl-serines in the brains of AD donors, particularly in carriers of *TREM2* risk variants, could reflect the presence of AD-associated damage signals in the form of stressed/apoptotic cells and damaged myelin.

## Introduction

Life expectancy has steadily increased in recent years, but with it brings greater prevalence of age-related conditions such as Alzheimer’s disease (AD), the most common cause of dementia. Dementia numbers are predicted to grow from 46.8M to 131.5M by 2050^1^ highlighting the urgent need for effective treatments. DNA variants linked to ∼84 genes are consistently associated with AD risk, of which ∼25% have highly enriched or specific expression in brain microglia (reviewed in^2^). Genes with immune or lipid function have been recognised for some time to be enriched in AD GWAS results^3^. One of these genes *TREM2*, codes for the Triggering receptor expressed on Myeloid Cells 2. Rare, but large-impact hypomorphic variants in *TREM2* significantly increase AD risk^4–7^ and related conditions^8–12^. Rare recessive *TREM2* mutations can also cause Nasu-Hakola disease characterised by fragility fractures, brain white matter changes and dementia in early adulthood^13^.

Most *TREM2* risk variants cluster in the extracellular Ig-like V-type domain impacting production (rs104894002, Q33X), expression or turnover of TREM2 at the cell surface (rs75932628, R47H)^14,15^, α-secretase cleavage of the extracellular ectodomain (sTREM2 production) (rs2234255, H157Y)^15–17^, shedding of sTREM2 (rs75932628, R47H)^18^ and/or ligand binding (rs75932628, R47H; rs143332484, R62H; rs2234253/rs2234258/rs2234256, T96K/W191X/L211P; rs2234255, H157Y)^19–21^.

TREM2 is a damage-response receptor expressed exclusively by myeloid cells including brain microglia^22^. It has a preference for binding anionic lipids, notably Phosphatidyl-serine (PS)^21,23–25^ a membrane signal on apoptotic cells, notably neuronal synapses in AD^26^. Other lipids reported to activate TREM2 include Phosphatidyl-ethanol (PE), Phosphatidyl-choline (PC), Phospholipids, and Sphingolipids such as Ceramides (Cer) and Sphingomyelins (SM). Damaged oligodendrocyte myelin and stressed or apoptotic cells expose sphingolipids in AD^19–21,23,24,27–31^. APOE and other apolipoproteins and Aβ oligomers also appear to be ligands for TREM2^20,32–36^.

TREM2-related pathologies include regional brain atrophy, myelin loss, swollen axons in white matter and changes the density and shape of microglia subsets^37,38^. TREM2 dysfunction prevents microglia from switching to a glycolytic state^39^ resulting in deficits in Aβ, myelin debris, apoptotic neuron and *E.coli* phagocytosis^15,40–43^. Results from analysis of very rare *TREM2* risk cases is limited. Nevertheless, non-hydroxy fatty acids of sulfatide (C16-C18) were found to be higher in Nasu-Hakola cortex compared to matched controls, while longer-chain (C24) fatty acids were lower^44^. Additionally, free fatty acids were increased^45^ while cholesterol and Ceramides in white matter were reduced^46^. Microglia from a cuprizone demyelinating TREM2-deficient mouse model were found to still sense and take up myelin debris but failed to efflux myelin cholesterol, resulting in cellular cholesteryl ester (CE) accumulation^25^. In that same study, shorter fatty acid Ceramides were elevated indicative of inflammation. Together, TREM2 deficiency appears to lead to broad lipid dysregulation.

In AD cases with a *TREM2* risk variant, plaques are more diffuse and nearby microglia have reduced membrane ruffling, and shorter but more numerous filopodia when stimulated with ATP or M-CSF^47^. Loss of TREM2 impairs actin ring formation and podosomes in osteoclasts, essential for phagocytosis linked to bone resorption^48^. PIP2 to PIP3 conversion at the cell membrane is essential for ‘sealing off’ the membrane edges during motility, endocytosis, exocytosis and phagocytosis through the cytoskeletal system. TREM2 signalling links to this pathway through Syk, MAPK, PIP2-PI3K-PIP3 Rac1 and Cdc42involving lipid species^31,49^.

Over-expression, activation or restoration of normal TREM2 function has largely beneficial outcomes in amyloidogenic mouse models^50–53^ while knock-out or haploinsufficiency exacerbates amyloid-associated pathologies^54–58^. These benefits are likely linked to improved phagocytosis of amyloid and clearance of damaged neurons by phagocytosis. It is noteworthy that amyloid plaques contain various lipids (Sphingolipids such as Ceramides, Phospholipids, Lysophospholipids as well as Cholesterol and Triglycerides) which may themselves influence pathogenesis^59–61^. Ceramides are potent inflammatory signalling molecules associated with AD in circulation^62^.

We hypothesize brain lipid dysregulation in AD reflects unresolved pathologies (damaged myelin, stressed/dying neurons and their knock-on impacts on lipid metabolism and membrane composition) exacerbated in people with a TREM2 risk variant where microglia fail to recognise and clear these pathologies effectively. Here, we sought to characterize networks or “modules” of highly connected lipids in post-mortem brain tissue from AD and control donors, and to identify key lipid “hubs,” i.e., highly connected lipids likely to play central roles in the functioning and regulation of these networks, associated with AD and impacted by TREM2. Specifically, we used a network approach used traditionally for genomic data, Weighted Gene Correlation Network Analysis (WGCNA), in post-mortem brain from two areas, the Hippocampus (HC) and the BA9 pre-association cortex. We investigated whether these networks and their “hubs” were altered in brain tissue from AD donors compared to control donors, and in brain tissue from AD carriers of rare *TREM2* risk variants (TREM2^var^) compared to controls and AD donors with no TREM2 risk variants (TREM2^wt^). Results from this study highlight lipid networks of highly correlated lipids and key “hubs” connected to AD pathology. It also provides insights into the role of lipids in TREM2-mediated activation of microglia in AD, which together highlight processes to target in future therapeutic strategies.

## Materials and Methods

### Study participants and samples

Informed consent for all brain donors was obtained according to the Declaration of Helsinki (1991) and protocols and procedures were approved by the relevant ethical committee and by each brain bank. Brain tissue was provided following project approval by the London Neurodegenerative Diseases Brain Bank (LNDBB). LNDBB subjects were approached in-life for written consent for brain banking, and all tissue donations were collected, stored and distributed following legal and ethical approval (Wales REC 3 favourable opinion 17 Sep 2013, REC number 08/MRE09/38+5; LBBND HTA license number 12293).

Pathological diagnosis was made according to established methods at the time of donation^63–67^. Where necessary for historical cases, retrospective diagnoses were made using current criteria. Post mortem human brain tissue, from the Hippocampus (HC) and BA9 pre-association cortex from n=60 brain donors were obtained for three groups: AD/TREM2^var^ (AD donors with a non-synonymous high AD risk DNA variant in TREM2), 20 AD/TREM2^wt^ (AD donors with AD-associated TREM2 variant) and 21 Control/TREM2^wt^ (tissue from age-matched donors absence of AD pathology or AD-associated TREM2 variant).

The TREM2^var^ group were selected from 19 donors: 9 donors identified by sequencing TREM2 Exon 2 in 631 AD and normal elderly control brain donors from the MRC London Neurodegenerative Diseases Brain Bank; 4 donors identified from a screen of 198 AD cases from The Netherlands Brain Bank (Royal Netherlands Academy of Arts and Sciences, Netherlands) and a further 6 donors described previously^4^ from the Queen Square Brain Bank for Neurological Disorders. From these, 14 had a pathologically confirmed diagnosis of AD while 5 were pathologically normal at death, although one displayed symptoms of mild cognitive impairment just prior to death and were subsequently assessed as Braak stage III at post-mortem. Consequently n=14 with AD were included in the AD/ TREM2^var^ group of which the majority were *TREM2* c.140G>A; p.R47H (rs75932628). One donor had a novel variant while the remaining variants have been described previously in AD cases (Table 1 and Table S1). In total, BA9 pre-association cortex was available for 55 donors while hippocampus (HC) was available for 47 donors (Table 1 and Table S1).

**Table 1.** Cohort Characteristics.

|  | BA9 (n=55) |  |  | EC (n=47) |  |  |
| --- | --- | --- | --- | --- | --- | --- |
| Diagnosis, n | AD<br>(n=34) | Control<br>(n=21) | P* | AD<br>(n=27) | Control<br>(n=20) | P* |
| Females/Males[% Female] | 20/34[59%] | 8/21[38.1%] | 0.224 | 16/27[59%] | 8/20[40%] | 0.312 |
| Age (years), mean (sd) | 74.4 (12.8) | 72.4 (12.6) | 0.566 | 76.5 (11.9) | 72.7 (12.9) | 0.300 |
| APOE ε4 alleles, n (0/1/2) | 6/21/7 | 17/3/1 | 2.25e-05 | 4/18/5 | 16/3/1 | 4.58e-05 |
| PMD (hrs), mean (sd) | 32.56 (22.5) | 32.81 (18.1) | 0.965 | 32.41 (21.66) | 32.4 (19.30) | 0.999 |
| TREM2<sup>var</sup>/total samples,<br>n[% TREM2<sup>var</sup>] | 14/34[28%] | 0/21[0%] | 2.0e-03 | 7/27[16%] | 0/20[0%] | 0.040 |
| TREM2<sup>var</sup>breakdown<br>p.R47H/p.T96K/p.D87N<br>/p.D39E/p.Q33X/p.G58A | 7/1/3/1/1/1 | 0/0/0/0/0/0 | NA | 4/1/1/0/1/0 | 0/0/0/0/0/0 | NA |

\**P*-values based on one-way analysis of variance for continuous variables or Chi square test for categorical variables

### Sample Preparation

Samples were randomized and lipids extracted based on an in-vial dual extraction (IVDE) protocol^68^. Briefly, 10μl of water was added to 50μl of the homogenate. Vials were then vortexed for 5 minutes, after which 250μl of methyl-tertiary butyl ether (MTBE) containing Tripentadecanoin (10μg/ml) and Heptadecanoic acid (10μg/ml) was added, and samples were again vortexed at room temperature for 60 minutes. Following the addition of a further 40μl of water containing 0.15mM ammonium, samples were centrifuged at 2500×g for 30 minutes at 4°C. This resulted in clear separation of an upper MTBE and lower aqueous phase.

### Data Acquisition

LC-MS Lipidomics analysis of the upper MTBE layer in positive mode was performed on a Waters Acquity ultra performance liquid chromatogram (UPLC) system coupled to a Waters Premier quadrupole time-of-flight (Q-Tof) mass spectrometer (Waters, Milford, MA, USA). Briefly, 5 μl of sample extract was injected onto an Agilent Poroshell 120 EC-C8 column (150mm × 2.1mm, 2.7μm). The gradient started at 80% mobile phase B increasing linearly to 96% B in 23 minutes and was held until 45 minutes then the gradient was increased to 100% by 46 minutes until 49 minutes. Initial conditions were restored in 2 minutes ahead of 7 minutes of column re-equilibration. Data were collected in the centroid mode over the mass range m/z 50–1000 with an acquisition time of 0.1 secondsper scan. Samples were analysed in a randomized order along with pooled brain samples (Quality control samples) after every eight injections.

### Data Pre-processing and Lipid Identification

All data was collected by Waters Xevo QTOF which used a MSe technique. Two collision energies were applied enabling data collection at two levels. The first level obtained data using 5 V of collision energy, the second level obtained data at a higher collision energy of 50 V. This can assist in structural elucidation and to simultaneously collect accurate mass of parent ion and fragmentation data. Following LC-MS analysis of samples, the MS raw data were transformed into mzXML format using msConvert (ProteoWizard). XCMS software package in R was then used to analyse the converted mzXML data files, which underwent preprocessing steps of peak picking and alignment processed, using a ‘centwave’ method which enables the deconvolution of closely eluting or slightly overlapping peaks. Data was normalised to total peak area, imputed and inverse variance transformed.

A total of 99 lipids were identified in the brain samples following our previous established protocol^69^. Identification was based on fragmentation patterns and comparison with lipid features from our in-house lipid library containing pure standards. Lipid species were examined in positive ion mode and the product ions of the[M+H]^+^ precursor was used to determine their acyl composition. As an example, the fragment ion 184.03Da corresponds to phosphocholine head group. Twenty-six Sphingolipids (16 Sphingomyelins and 10 Ceramides) were identified with a parent molecular ion in the form[M-H2O+H]^+^ and the fragment ion 264.26Da which corresponds to a Sphingosine base chain. We named the fatty acid chains as very long (VLCH), longer (LCH) or shorter (SCH), but these names are only for reference in this manuscript, as there isn’t a single universal standard specifically for lipid naming based on chain length.

## Statistical Analysis

### Data Quality Control

Lipid features missing in ≥20% of donors, and donors with ≥20% missing features were excluded from further analyses. For the remaining data, missing data points (n=4) were imputed using k-nearest neighbours (knn, k=10) (“impute”) and the dataset was subsequently inverse-normal-transformed.

### Preliminary associations between lipids, brain regions and covariables

In preliminary analyses, we investigated the association of each lipid with sex, age (at death), number of APOEε4 alleles and post-mortem delay, combining the two brain regions (Supplementary Table 1). Linear model analysis was performed with lipids as outcomes using generalized least squares (‘gls’ function in nlme R package^70^), which allows for a fully unstructured residual variance-covariance matrix. Analyses were initially performed separately for each covariable, adjusting for post-mortem delay with sex, age (at death), number of APOEε4 alleles as predictors. An interaction term between brain region and each predictor, was included in each model to investigate possible brain region-specific associations. In the presence of an interaction, linear regression analyses were repeated, stratifying for each brain region. To correct for multiple testing and the high correlation between some lipids (Supplementary Figure 1), we set a metabolome-wide statistical significance threshold of *p* < 0.001; the *p* < 0.05 significance level was divided by the number of principal components (n=45) that explained over 95% of variation in the lipidomic data.

## Weighted Gene[Lipid] Co-expression Analysis

### Network construction

To define lipid networks within the BA9 pre-association cortex and Hippocampus, we applied WGCNA to the lipid data for each brain region. Lipids were first adjusted for sex, age (at death) and post-mortem delay, separately in each region, and the standardized residuals were used for subsequent analyses. Next, the standardized connectivity (*Z*.*k*) for each sample was computed to identify outliers (Z>|4|). A pairwise correlation matrix using biweight midcorrelations between all lipids was then derived. From this, a weighted, signed adjacency matrix was constructed by raising correlations to a soft thresholding power of 12 for both modules, chosen to meet a scale-free topology threshold of ≥0.8, while maximizing mean connectivity.

Subsequently, the adjacency matrix was transformed into a topological overlap matrix (TOM), representing the network connectivity of lipids. Lipids were then hierarchically clustered into a dendrogram using an average linkage method based on their dissimilarity (1−TOM), and the dendrogram was cut using a dynamic hybrid tree cutting algorithm parameters—minModuleSize = 12 (to allow for smaller modules to be constructed), and mergeHeight = 0.25 (default)).

The resulting modules or groups of co-expressed lipids were used to calculate module eigengenes (MEs; or the 1st principal component of the module) for all modules. The ‘grey’ module comprised lipids that were not assigned to any particular module and was therefore dropped from further analyses.

Lipids that are highly connected to their module (termed “hub” lipids) are likely to be functionally important. To identify hubs lipids in each module, the associations between lipids and their assigned module (module membership; kME) were calculated using correlations between lipids and module eigenvalues. Lipids with a kME > 0.70 in each brain region were defined as hubs.

### Module preservation between BA9 and Hippocampus

To investigate whether modules between the two brain regions showed similar coexpression/connectivity and were thus preserved, we utilized the WGCNA “modulePreservation” function. Module preservation and robustness was summarised by reporting Z summary scores, a composite measure of 4 statistics related to density and 3 statistics related to connectivity. The module preservation analysis was applied twice assigning one as the reference dataset and the other as the test dataset. Summary values between 2 and 10 are considered to be moderately preserved (reproducible), while those below 2 are considered not preserved, and those above 10 are considered strongly preserved^71^.

### Associations between lipid modules, AD and TREM2 in each brain region

Following WGCNA analyses, we sought to investigate the association between lipid modules in each brain region and 1) Post-mortem AD diagnosis (i.e. combining AD/TREM2^var^ and AD/TREM2^WT^) *vs* Control/TREM2^WT^ donors. To probe whether any associations of brain lipid modules with AD diagnosis were driven by genetic variation at the TREM2 locus we then compared brain lipid module levels in 2) AD/TREM2^WT^ brain donors *vs* Control/TREM2^WT^ donors, 3) AD/TREM2^var^ *vs* Control/TREM2^WT^ donors, and finally,4) TREM2^var^ *vs* TREM2^WT^ in AD donors only (i.e. AD/TREM2^var^ *vs* AD/TREM2^WT^). Analyses were performed for each brain region separately by regressing the residualised lipid modules generated through WGCNA against the outcomes using linear regression analyses and further adjusting for the number of APOEε4 alleles. To investigate the association of APOEε4 with lipid modules, separate regression analyses between each module and the number of APOEε4 alleles were additionally fitted. For all module analyses, a Bonferroni-corrected significance threshold was set at p< 0.05/ Number of modules for each brain region(4) = 0.0125.

### Associations between hub lipids, AD and TREM2

We finally investigated the associations with hub lipids (kME>0.70) in modules associated with AD and/or TREM2 in either brain region by performing a linear model analysis using generalized least squares, as described above, and by adjusting for the number of APOEε4 alleles. To further identify brain region-specific associations, an interaction between diagnosis and variant status and brain region was included in each GLS model. To avoid multiple testing issues, an interaction was considered between brain region and AD vs Controls, and between brain region and AD/TREM2^WT^ vs AD/TREM2^var^, as these would capture brain region AD-specific or TREM2-specific effects. In the presence of an interaction (p<0.05), linear regression analyses were repeated, stratifying for each brain region. For all hub analyses, a Bonferroni-corrected significance threshold was set at p< 0.05/ 31 (Number of hubs) = 0.0016.

Analyses were conducted in RStudio (R version 3.4.2).

## Results

### Donor and sample characteristics

The demographic characteristics of the cohort for each brain region are described in Table 1. Overall, BA9 pre-association cortex tissue was available for 55 donors while hippocampus tissue was available for 47 donors. Although AD donors were generally older, and more likely to be female, there was no statistical difference between age at death, post-mortem delay. The ratio of men to women was similar across groups (p=>0.05) (Table 1). Of the 99 annotated lipids 10 were Ceramides (CER), 16 were Sphingomyelins (SM), 6 were Phosphatidic acids (PA), 22 were Phopshatidy-lcholines (PC, including 4 Lyso-phospholipids), 2 were Phosphatidyl-glycerols (PG), 6 were Phosphatidyl-inositols (PI), 13 were Phosphatidyl-serines (PS), 12 were Phosphatidyl-ethanolamines and 12 were Triglycerides (TG). Overall, PS levels were higher compared to other lipids (Supplementary Figure 2).

### Associations of lipids with age at death, sex, APOEε4 genotype and post-mortem delay

Most lipids had different levels in BA9 compared to hippocampus. After controlling for multiple testing, 55 out of 99 lipids differed between the two brain regions, 32 of which were lower in the hippocampus and 23 higher in the hippocampus compared to BA9 (Supplementary Table 1).

Following correction for multiple testing, only one lipid, Phosphatidyl-choline (PC(38:2)), showed an association with the number of APOEε4 alleles (beta=-0.49, 95% CI = –0.78 – – 0.21, p=0.0009). No lipids were associated with sex, age at death or post-mortem delay. Overall, the association of lipids with the covariates were consistent between the two brain regions, in terms of direction of effects, with no interactions present after multiple testing correction (Supplementary Table 1).

### Network analyses

WGCNA analysis identified four modules in each brain region, comprising 16–22 lipids in the BA9 cortex and 14-26 lipids in the Hippocampus. A grey module (lipids not assigned to one of the four modules) included the remaining 22 and 17 lipids, respectively.

The four modules identified in BA9 were the turquoise module, comprising mainly of Longer Chain Ceramides and SMs, and Medium Chain Phosphatidyl-choline (PC) lipid species; the yellow module comprising mainly Very Long Chain Phospholipids; the brown module comprising Very Long Chain SMs and Phospholipids, and the blue module comprising mainly Triglycerides (TGs).

In the Hippocampus, the blue module (corresponding to the BA9 turquoise module) comprised mainly Longer Chain Ceramides and SMs, and medium chain PC lipid species; the yellow module (equivalent to the BA9 yellow module) comprising mainly Very Long Chain Phospholipids; the turquoise module (corresponding to the BA9 brown and yellow modules) comprising mainly Very Long Chain Phospholipids and Sphingolipids; and the brown module (corresponding to the BA9 blue module) comprising mainly TGs. Module preservation analyses indicated that all four lipid modules showed medium-to-high preservation and reproducibility between the two brain regions (Supplementary Figure 3).

## Module-level regression analyses

### BA9

In linear regression analyses, AD donors had higher turquoise module levels and lower yellow module levels compared to control donors after adjustment for multiple testing (beta= 1.003, 95% CI =0.42 –1.59, p=0.001 and beta= –0.821, 95% CI =-1.44 – –0.20, p =0.011, respectively) (Figure 2 and Supplementary Table 2). We next sought to explore whether any associations in lipid module levels observed between AD and control brains were driven by TREM2 risk variants, by separating AD/ TREM2^WT^ and AD/TREM2^var^ donors. For the turquoise module, we observed additive AD and TREM2 effects, whereby levels of lipids in the turquoise module were increased on average by 0.76 SD in AD/ TREM2^WT^ donors compared to controls (95% CI = 0.13 – 1.37, p =0.019) and by 1.35 in AD/ TREM2^var^ donors compared to controls (95% CI = 0.69 – 2.01, p = 1.467* 10^−4^), although only the association between AD/ TREM2^var^ donors and controls survived multiple testing correction. Comparing BA9 turquoise module levels between AD/ TREM2^WT^ donors and AD/ TREM2^var^ donors highlighted a modest increase in TREM2 carriers (beta=0.601, 95% CI= 0.008 – 1.19, p =0.047). On the other hand, the decrease in yellow module levels in AD/ TREM2^WT^ donors relative to controls was similar to that observed in AD/ TREM2^var^ donors compared to controls (beta=-0.834, 95% CI –1.52 to –0.145, p =0.019 and beta=-0.03, 95% CI –1.54 to – 0.07 p =0.032 respectively), highlighting no additional effect of TREM2 on lipid levels in the yellow module (TREM2-independent associations). None of these associations were significant after correction for multiple testing (Figure 2 and Supplementary Table 2).

**Figure 1.**
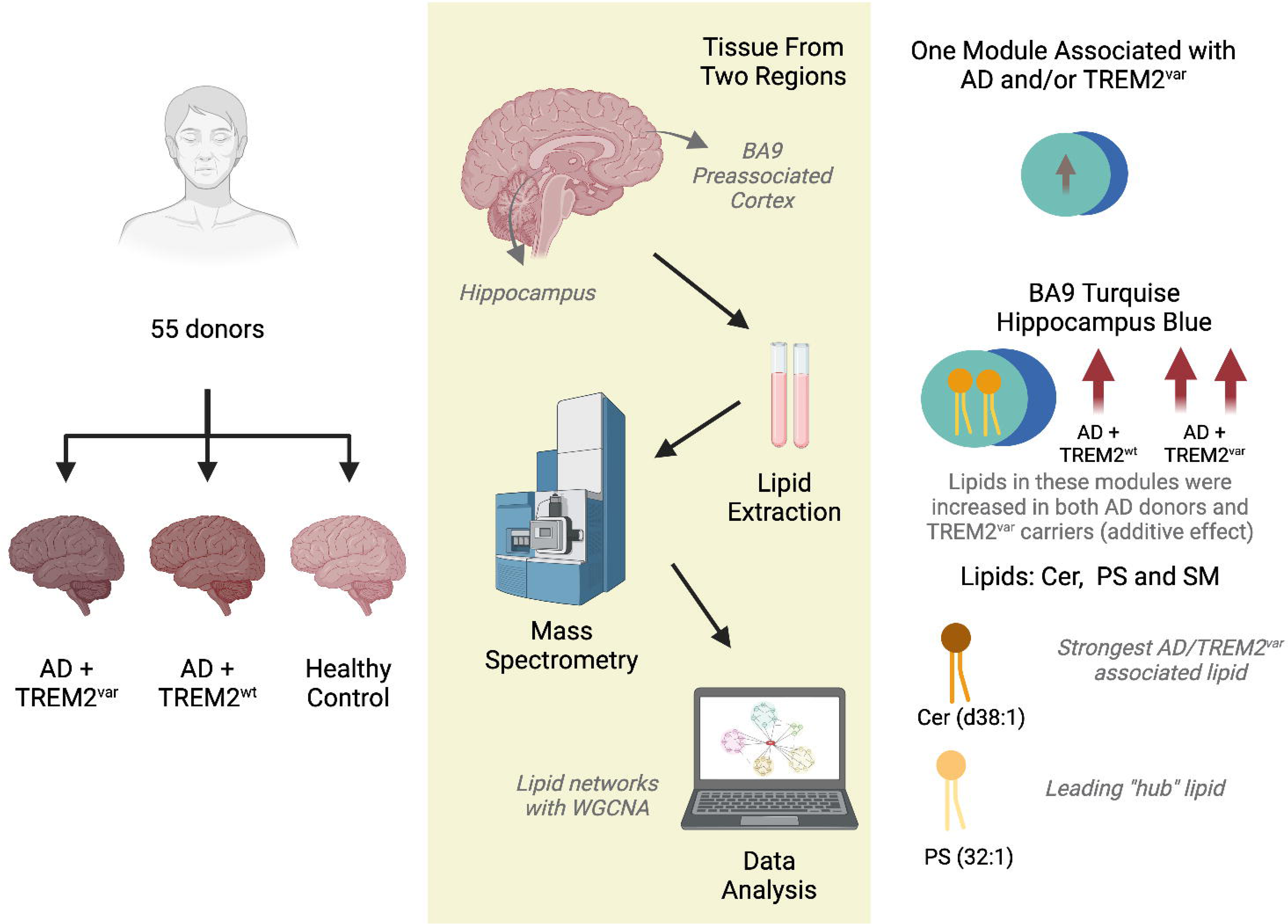
Diagramatic study flow.

**Figure 2.**
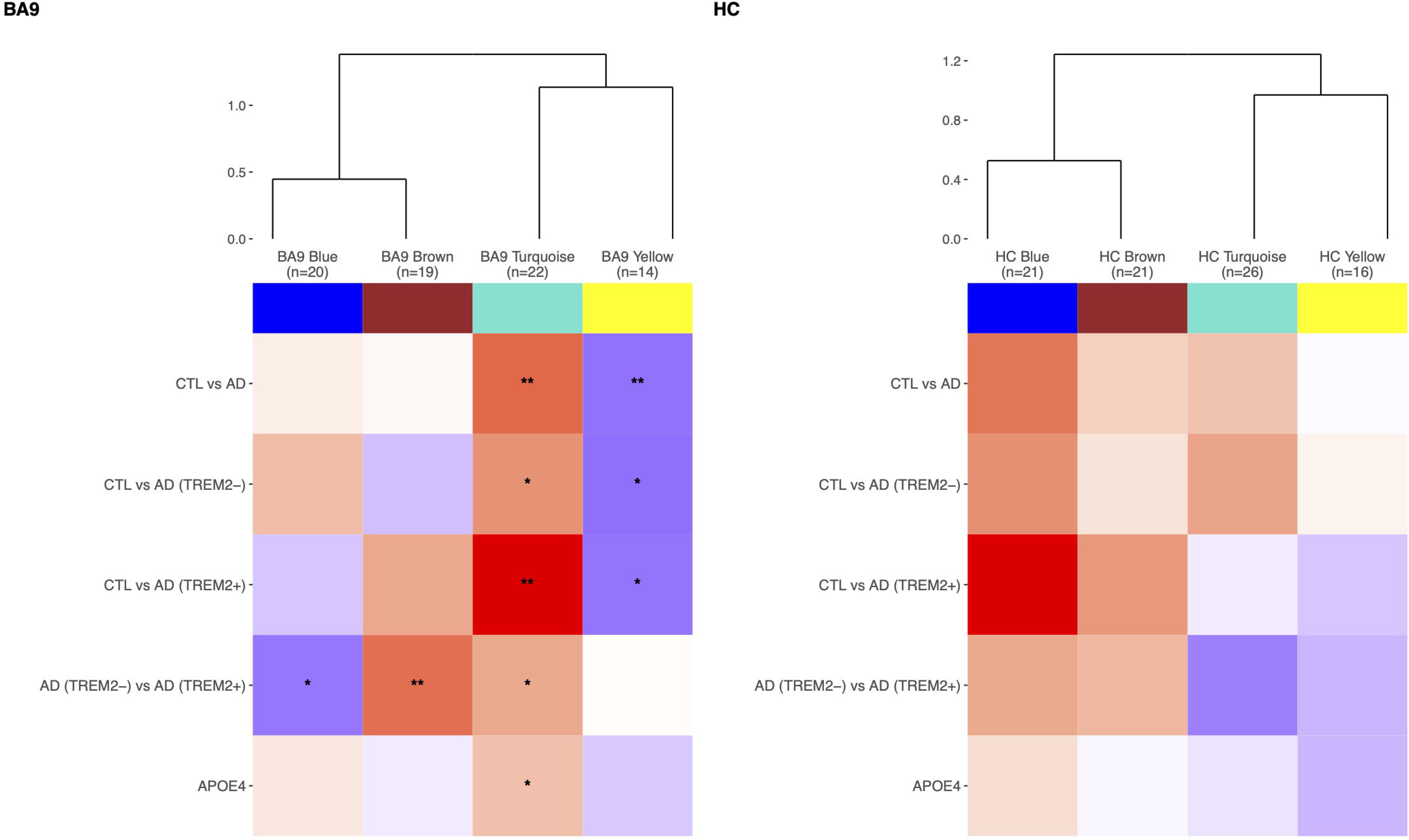
Associations of lipid modules with post-mortem diagnosis and TREM2 status. a) Heatmaps showing the relationships of BA9 and Hippocampus (HC) modules with 1) Post-mortem AD diagnosis (i.e. AD/TREM2^var^ and AD/TREM2^WT^ combined vs Control/TREM2^WT^ donors), 2) AD/TREM2^WT^ brain donors vs Control/TREM2^WT^ donors, 3) AD/TREM2^var^ vs Control/TREM2^WT^ donors, 4) TREM2^var^ vs TREM2^WT^ in AD donors, and 4) Number of APOEε4 alleles. The number of lipids in each module is shown underneath each module name. *p<0.05, **p<0.00125. The colour of the tiles reflects the strength of the association (beta) with blue tiles indicating a decrease and red tiles indicating an increase in lipid levels in AD donors vs controls, in TREM2+ vs TREM2-carriers and in donors with a higher number of APOEε4 alleles. CTL=controls, AD=Alzheimer’s Disease, APOE4=number of APOEε4 alleles. *The BA9 blue module corresponds to the Hippocampus brown module; the BA9 brown module corresponds to the Hippocampus turquoise module; the BA9 turquoise module corresponds to the Hippocampus blue module; the BA9 yellow module corresponds to the Hippocampus yellow and turquoise modules*.

We also observed higher brown module lipid levels and lower blue module levels in AD/ TREM2^WT^ carriers vs AD/ TREM2^var^ (AD-independent associations), although only the association with the brown module survived multiple testing (beta=0.958, 95% CI= 0.29 – 1.64, p =0.006). Finally, we observed a nominal positive association between the turquoise module and the number of APOEε4 alleles testing (beta=0.443, 95% CI= 0.07 – 0.81, p =0.020).

### Hippocampus

Linear regression analyses revealed that the direction of effect of the observed associations in the Hippocampus was similar to those observed in BA9, particularly for the Hippocampus blue module (corresponding to the BA9 turquoise module). However, the strength of the associations in the hippocampus was weaker to that in BA9 with no nominal associations observed (Figure 2 and Supplementary Table 2).

### Association of hub lipids with AD and TREM2

We next sought to identify associations between hub lipids that were highly connected (kME>0.70) in modules associated with AD and/or TREM2 after correction for multiple testing i.e. the BA9 turquoise, brown and yellow modules. The candidate 31 hubs included 12 lipids from the BA9 turquoise module, 12 lipids from the BA9 yellow module and 7 lipids from the BA9 brown module. The top hub for the BA9 turquoise module was Phosphatidyl-serine (PS) (PS(32:1)), followed by longer chain Ceramides and SM, and Phosphatidyl-ethalonamines; the key hub for the BA9 yellow module was Phosphatic Acid (PA 36:1) followed by other phospholipids; and the key hub for the BA9 brown module was SM (SM d44:2), followed by other SMs. Since all the modules showed medium-high preservation between the two brain regions, joint brain-area analyses were performed for each lipid using generalised least squares (GLS) and setting a Bonferroni-corrected significance threshold at p< 0.05/ 31= 0.0016.

Overall, ten hub lipids were found to be associated with AD and/or TREM2 status after multiple testing correction (Figure 3, Supplementary Table 3). All ten lipids were in the BA9 turquoise module (HC blue module) that consisted of medium chain Phospholipids and longer chain Sphingolipids.

**Figure 3.**
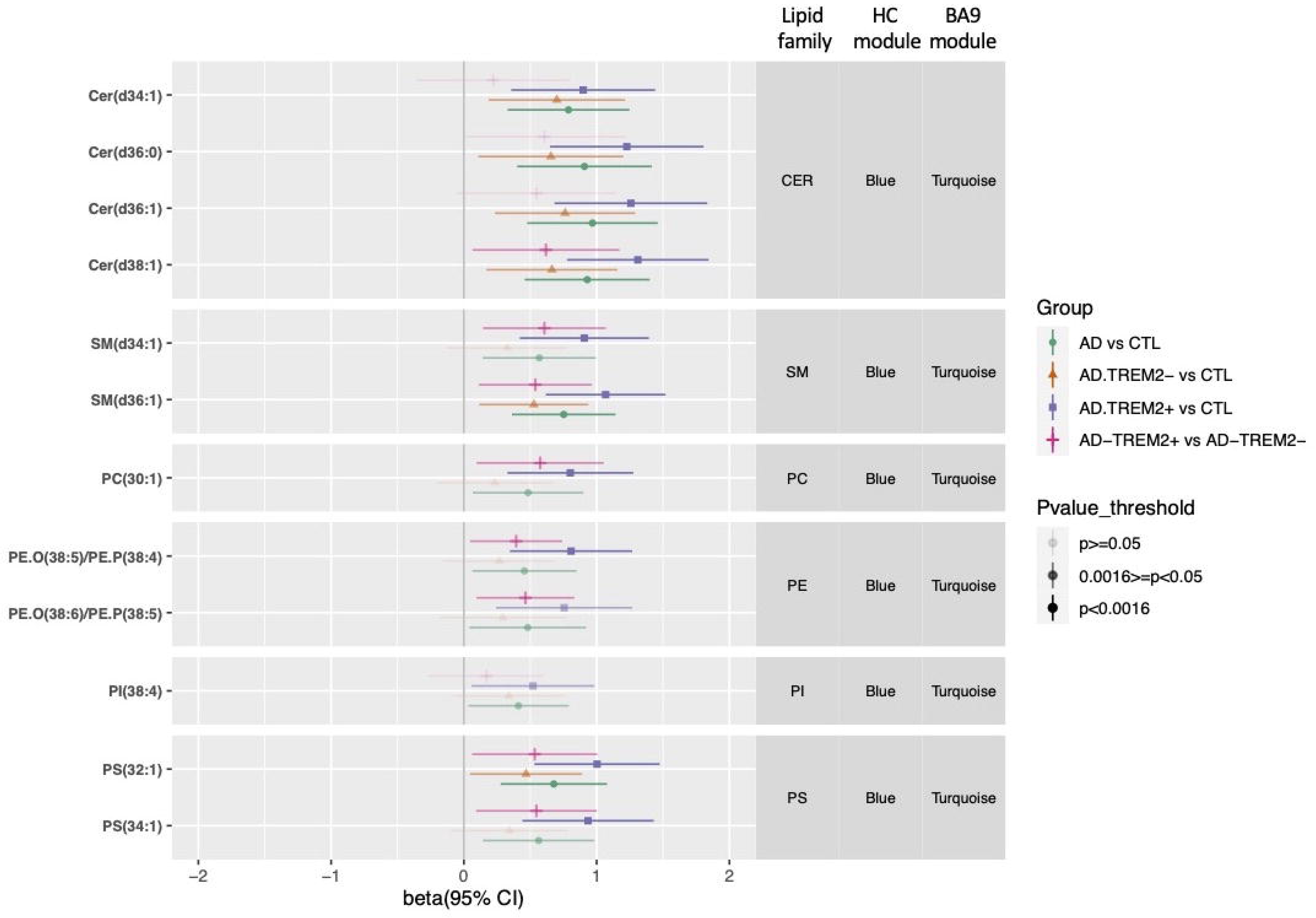
Association of hub lipids with Post-mortem AD diagnosis and with TREM2 status. 1) Post-mortem AD diagnosis (i.e. AD/TREM2^var^ and AD/TREM2^WT^ combined vs Control/TREM2^WT^ donors), 2) AD/TREM2^WT^ vs Control/TREM2^WT^ donors, 3) AD/TREM2^var^ vs Control/TREM2^WT^ donors, 4) TREM2^var^ vs TREM2^WT^ AD donors, (BA9 and Hippocampus combined) using generalised least squares and adjusting for sex, age (at death), post-mortem delay and APOE4 genotype. The columns on the right of the plot indicate the lipid family and module each lipid is assigned to, in each brain region. Lipid levels are standardised to a mean of 0 and SD of 1. Bonferroni adjusted threshold p<0.00146. CTL=control donors; AD=Alzheimer’s disease donors; CER=Ceramide; SM=Sphingomyelin; PA=Phosphatidic acid, PC=Phosphatidyl-choline, PI=Phosphatidyl-inositol; PS=Phosphatidyl-serine; PE=Phosphatidyl-ethanolamine.

Notably, six of the lipids were increased in AD compared to controls after adjustment for multiple testing. The strongest relationship was with Ceramide (Cer(d38:1); beta=0.929, 95% CI 0.46 – 1.40, p= 1.68E-04) (Figures 3 and 4), with the remaining associations with three Ceramides, one Sphingomyelin (SM(d36:1)) and with the BA9 turquoise and HC blue modules key hub Phopsphatidyl-serine (PS(32:1)) (beta=0.677, 95% CI 0.28-1.08, p= 1.14E-03).

**Figure 4.** Levels of Cer(d38:1) and PS(32:1) in AD and Controls, and in TREM2^WT^ and TREM2^var^carriers in BA9 and the Hippocampus. Cer(d38:1) was the top turquoise module lipid associated with AD (combined AD/TREM2^var^ and AD/TREM2^WT^) vs Control/TREM2^WT^ donors and with AD/TREM2^var^ vs Control/TREM2^WT^ donors, and PS(32:1) was the top “hub” lipid in the turquoise module (i.e. the hub lipid more closely associated to its module).

We then sought to explore whether any associations between the six hub lipids and AD were driven by TREM2 risk genotypes. Similarly to module level analyses, GLS analyses for both brain regions highlighted an increase in all BA9 turquoise / HC blue lipid levels ranging from ∼0.47 SD to ∼0.76 SD between AD/ TREM2^WT^ donors and controls, and an increase in lipid levels ranging from ∼0.80 SD to ∼1.31 SD between AD/ TREM2^var^ donors and controls, highlighting on average an increase of ∼0.5 SD in lipids levels between AD/ TREM2^WT^ and AD/ TREM2^var^. Although the associations between AD/ TREM2^WT^ and controls were nominal (p<0.05), all six associations between AD/ TREM2^var^ and controls were significant after correction for multiple testing (Figure 3 and Supplementary Table 3). The strongest association was again with Cer(d38:1)) (beta=1.310, 95% CI 0.78 – 1.84, p= 4.35E-06). We also observed four additional lipid hubs, all belonging to the BA9 turquoise / HC blue modules, that were increased in AD/ TREM2^var^ vs controls after correction for multiple testing, and which had shown only nominal associations with AD status (Figure 3, Supplementary Table 3). These results highlight additive AD and TREM2 effects for most lipids in the BA9 turquoise and HC blue modules. Finally, we observed some associations with hub lipids in other modules suggesting TREM2-independent and AD-independent effects and reflecting the results observed at module level. None of these associations remained significant after adjustment for multiple testing (Supplementary Table 3).

### Brain region specific association of lipids with AD and TREM2 variants

Overall, we found associations between brain lipids and end-stage AD with or without a TREM2 mutation to be consistent between the two brain regions, particularly for the BA9 turquoise /hippocampus blue modules, with overall stronger effects observed for BA9.

There was weak evidence for brain-region-specific associations between TREM2 and lipids belonging to the BA9 yellow module, where hub Phospholipids were higher in AD donors compared to controls, with no further increase in TREM2 carriers (TREM2-independent associations) (interaction Pvalue <0.5) (Supplementary Table 3). We also observed that the levels of hub lipids belonging to the BA9 brown module, particularly SMs, were increased in AD/ TREM2^var^ compared to AD/ TREM2^WT^ (AD-independent associations) (interaction Pvalue <0.5), although these relationships did not survive multiple testing correction (Supplementary Table 3).

## Discussion

Here, we characterized lipid networks in 102 post-mortem brain samples from 55 AD and control donors, identifying key lipids associated with AD and impacted by dysfunctional TREM2. TREM2 is an established lipid receptor uniquely expressed by microglia in the brain. It is also a risk gene for AD^19–21,23,27–31^. Four modules consisting of highly correlated lipids were well-preserved between the BA9 pre-association cortex, and hippocampus. Lipid levels in the BA9 “turquoise” module (hippocampus “blue” module) were significantly elevated in post-mortem tissue form AD donors compared to controls. Levels were even higher in AD donors who additionally had a loss-of-function TREM2 AD risk variant. The BA9 “turquoise”/hippocampus “blue” module was enriched in longer chain Sphingolipids (Ceramides and Sphingomyelins), and Phospholipids including Phosphatidyl-serines. Overall, the strongest associations were observed with Ceramides, whereas the key hub lipid i.e. lipid with the highest number of connections to other lipids in this module, was Phosphatidyl-serine PS(32:1) in BA9 and hippocampus. These findings further implicate TREM2 in lipid regulation.

Phosphatidyl-serine a major component of cell membranes, is normally confined to the inner cytoplasmic leaflet by transporter proteins flippase and floppase. However, when these enzymes are inactivated during cell death, it becomes exposed on the outer plasma membrane ^72^ becoming a phagocytic signal for macrophages including microglia^73,74^. Local exposure of Phosphatidyl-serine on neurons can impact spine engulfment and thus neural activity^26^.

Phosphatidyl-serine can also impact the structure and function of membrane nanodomains, including cholesterol linked functions^75^. Phosphatidyl-serine is an established ligand for the TREM2 receptor^21,23–25^. Our data shows that levels of Phosphatidyl-serine were even higher in *TREM2* AD risk variant carriers suggesting a failure by microglia to recognise and clear dysfunctional neurons. Senescent AD microglia are considered poor at degrading aggregate pathologies, leading to aggregate spreading and seeding ^76–79^. Nevertheless, they appear very capable of phagocytosing soma, nucleus and apical dendrites of neuronal corpses and together with astrocytes can fully degrade neurons^80^. In the course of clearing or partially clearing neuronal corpses in AD brain, insoluble extracellular neurofibrillary “ghost tangles” are left where neurons once resided ^81^. This suggests cell membranes containing Phosphatidyl-serine can be cleared in AD, although whether microglia partially or fully clear neurons in AD in those with a TREM2 risk variant has not been investigated. In microglia where TREM2 function is compromised, they fail to switch from homeostatic OXPHOS to activated glycolysis^39^ necessary to drive phagocytic function^23,82^ including of myelin debris^24^. It would be interesting to evaluate the number of “ghost tangles” in TREM2 cases and establish whether membrane remnants remain at these sites to better understand the contribution of TREM2 to neuronal clearance by microglia. A failure to remove apoptotic neurons in AD could lead to the persistence of damaging inflammatory signals from neurons such as HMGB1, DNA and IL-1α which in turn would exacerbate AD pathologies further^83–86^. It will also be important to evaluate the function of other microglia Phosphatidyl-serine receptors in the absence of TREM2 function. MERTK & Axl, integrin (ITGAV, ITGB3, ITGB5), HAVCR1 & TIMD4, ADGRB1, STAB1 (bridged by C1q), CD300f and LRP1 (bridged by Crt, C1q) are all highly or uniquely expressed by microglia^87^ and may become preferential Phosphatidyl-serine receptors in the absence of TREM2, altering the function of microglia in AD.

Phosphatidyl-serine can exist with differing acyl length and saturation^88^. PS(32:1) was readily detectable in the two brain areas we investigated and showed the strongest association with AD and TREM2, although PS(34:2) was the most abundant in our samples. There were 13 PS molecules detected in our study, four of which belonged to the BA9 “turquoise” module / hippocampus “blue” module, where levels were increased in AD and TREM2. It is unknown if there is a preference for different PS species by TREM2 and other PS receptors or why certain PS species are selectively elevated in AD. Phosphatidyl-serine is not only present in neurons, but is also highly abundant in myelin^89^ and other cell types (astrocytes and microglia)^90^, and these could also be a source of the elevated levels we observed in AD and in those with dysfunctional TREM2. Myelin levels could be particularly relevant in light of the raised Ceramide levels we also observed in AD, particularly in those with a TREM2 risk variant and our finding that TREM2 is central to the co-ordination of oligodendrocyte and microglia genes in AD^91^.

The strongest association with AD and TREM2 dysfunction in our samples was with longer chain Ceramides. These lipids were important hubs within the BA9 “turquoise” and hippocampus “blue” module, with their kME being slightly lower than that of Phosphatidyl-serines. Ceramides (galactosylceramide and sulfatide species) are synthesised predominantly by oligodendrocytes in the brain where they are the dominant lipid component of myelin^89,90,92^. Ceramide species accumulate in AD brains^93^ and leukodystrophies when dysfunctional enzymes required for their metabolism become dysfunctional. This leads to accumulation of intermediate lipid species, notably in microglia lysosomes^89,94^. Ceramides are metabolised to Sphingomyelin and vice versa, hence they can be a breakdown product of Sphingomyelin metabolism^93^. This perhaps explains why Sphingomyelin and Ceramide species were highly correlated and both appeared in the same modules in our study. Together these results, suggest TREM2 mediated microglia function is linked to oligodendrocyte function in AD.

Much effort has consolidated our knowledge of lipid intermediates and the enzymes responsible for their biosynthesis. Acyl chain length and saturation can impact lipid function and intermediates reflect sequential biosynthesis via *de novo* or salvage β-oxidation pathways. Typically, fatty acid chains are categorised as short-(<C5 carbons), medium-(C8-C13 carbons), longer-(C14-C20 carbons) and very long (>C20 carbons), although these categories vary by lipid group^95,96^ although there isn’t an agreed single universal standard for lipid naming based on chain length. In our study, it was longer chain and not very long chain Ceramides which had increased levels in AD brain and in those with a TREM2 risk variant (notably Cer (d38.1)). Ceramides, play important roles in membrane integrity, and can had proinflammatory and apoptotic function. We and others have previously found longer chain Ceramides and SMs to be elevated in the plasma of AD/dementia patients and are associated with hippocampal atrophy^62,96–98^. Consistent with our findings, others have also reported elevated levels in AD brain^99,100^. A recent study has further reported that Ceramides mediate the effect of known AD genetic factors such as ABC7 in AD^101^. Ceramide species like the ones reported here have been also implicated in cardiometabolic and cerebrovascular diseases and we have further demonstrated associations with 6-year cardiovascular risk and all-cause mortality in a Type-1-Diabetes cohort^102^. As both Ceramides and SMs increase the risk of cardiovascular disease and insulin resistance it has been also suggested that increased cardiometabolic risk could be mediating the association of ceramides with dementia risk^96^.

Finally, higher levels of Ceramides and SMs have been observed in a cuprizone model of myelin damage in mice in which TREM2 is absent^25^. In fact, sulfatides, which are synthesized from galactosylceramides, which in turn are derived from ceramides, are strong TREM2 activators^103^. We were unable to distinguish sulfatide species in our anlaysis.

In Nau-Hakola disease where there is complete loss of TREM2 function or its adapter DAP12, lysosomal function in microglia is impaired suggesting the possibility that the increased lipids we observed may be accumulating in microglia thus impacting there response to AD damage^104^. It will be interesting to establish if microglia from TREM2 AD patients accumulate lipids^105^, as was recently shown in AD patients carrying APOEε4^106^. Amyloid deposits also contain lipids including Ceramides^107^ and this could be an additional source of the elevated Ceramides we measured. Overall, our results suggest a failure by microglia to recognise damage lipid signals and successfully mobilise to clear damaged myelin and cells in AD leading to accumulation of specific lipids.

Ceramides can be potent bioactive molecules. High levels of ceramide can cause a myriad of changes culminating in increased damaging reactive oxygen species, through blockade of the respiratory chain, mitophagy and activation of apoptotic factors linked to axonal degeneration and cell death^93,108,109^. Furthermore, altered membrane lipid composition, particularly galactoceramides, can impact APP processing and amyloidogenic Aβ production^108,110^ and this along with increased TNF-α can impact enzymes that metabolise lipids including those required for Sphingomyelin to Ceramide metabolism, as well as factors linked to apoptosis of oligodendrocytes^111–113^. In microglia without fully functioning TREM2, a failure to recognise and resolve lipid signals adequately would exacerbate a cycle of damage.

We also found weak evidence for brain region-specific lipid changes. For example, very long chain SMs and Phospholipids in the BA9 brown module were increased in AD/TREM2^var^ compared to AD/TREM2^WT^ donors (AD-independent associations). Hubs within this module also showed nominal associations with TREM2, but only inBA9. Similarly, the BA9 yellow module, enriched in very long chain Phospholipids, was decreased in AD compared to control donors, with no further decrease observed in TREM2 carriers (TREM2-indenepdent association). Levels of two hub Phospholipids were increased in AD donors compared to controls in BA9 only, after adjustment for multiple testing (Supplementary Table 3). These findings warrant further investigation.

Finally, we found that Phosphatidyl-choline, PC(38:2) levels were lower in APOEε4 carriers and also in BA9 in AD donors, particularly those with a TREM2 risk variant (Supplementary Table 3). PC(38:2) showed borderline association with the BA9 yellow module (kME=0.694) and was a hub for the equivalent Hippocampus yellow module (kME=0.920). The relationship therefore between TREM2, APOE and PC(38:2) warrants further investigation. Phosphatidyl-cholines are primarily found in cell membranes including the monolayer encapsulating lipid droplets, a reservoir of neutral lipids and cholesterol esters that swell in AD microglia, particularly in the absence of TREM2 and in high risk APOEε4 carriers^106,114–117^. Their role extends beyond structural integrity, as they are an essential component of lipoproteins (VLDL/LDL/HDL) that facilitate transport of triacylglycerols/cholesterol to/from cells, a tightly regulated process, with APOE-containing LDL– and HDL-cholesterol particularly implicated in AD^118–120^. The decrease in brain Phosphatidyl-choline species in AD compared to control donors reflects reports by us and others in plasma ^62,121–123^. Additionally, Phosphatidyl-choline species like the ones reported here have been positively associated with hippocampal brain volume and negatively with disease progression^62^, as well as with CSF Aβ1–42^124^.

Although dysregulation of Sphingolipids and Glycerophospholipids in blood and brain has been previously reported by us and others^62,121–124^, this is one of the first studies to comprehensively investigate changes in brain lipid levels in AD donors carrying rare TREM2 risk variants and the first to use two AD-affected brain regions. Others have investigated impacts of TREM2 on brain gene expression^91^ and metabolites^125^ with findings implicating microglia, oligodendrocyte and endothelial genes, notably those involved in complement and Fcγ receptor function, microglia-associated ribosomal genes and oligodendrocyte genes, particularly proteosomal subunits and amino acid and sphingolipid metabolism and vitamin pathways, respectively. Recently Novotny *et al.*,^125^ compared metabolite levels in donated brain tissue from sporadic AD, familial AD and AD/TREM donors compared to control donors. They reported reduced levels of beta-citrylglutamate in AD and AD/TREM2 brains compared to controls, possibly related to lower energy metabolism, as well as reduced *α*-tocopherol and CDP-ethanolamine in AD/TREM2 brains compared to controls brains, and reduced ergothionine and N-acetylputrescine levels in AD compared to control brains. Future analyses should investigate associations between the lipids we identified, gene expression and protein levels to better understand the important molecular processes.

Our study has some limitations. The sample size was modest, particularly for the AD/TREM2^var^ group. However, the majority of donors had brain tissue from both areas for which findings were consistent. Our analyses are also limited to end-stage disease changes. The use of bulk tissue may obscure cell-type-specific lipid alterations. Future studies should focus on analyzing lipidomics for TREM2 carriers in larger cohorts investigate cell type changes and expand analyses to blood and CSF. It is also important to examine lipid changes in brains from ethnically diverse donors to generalize findings.

Overall, our results are consistent with TREM2 dysfunction interfering with the recognition and clearance of unhealthy neuronal cells, damaged myelin and lipid-containing amyloid.

These findings could have great translational potential as they highlight processes to target in future therapeutic strategies.

## Data availability statement

Data is available upon reasonable request from the corresponding authorsfs in collaboration with the authors.

## Supplementary material

Supplementary material is available online.

