## Supplementary Figures for "Alterations in the Brain Lipidome of Alzheimer’s Disease Donors with Rare TREM2 Risk Variants"

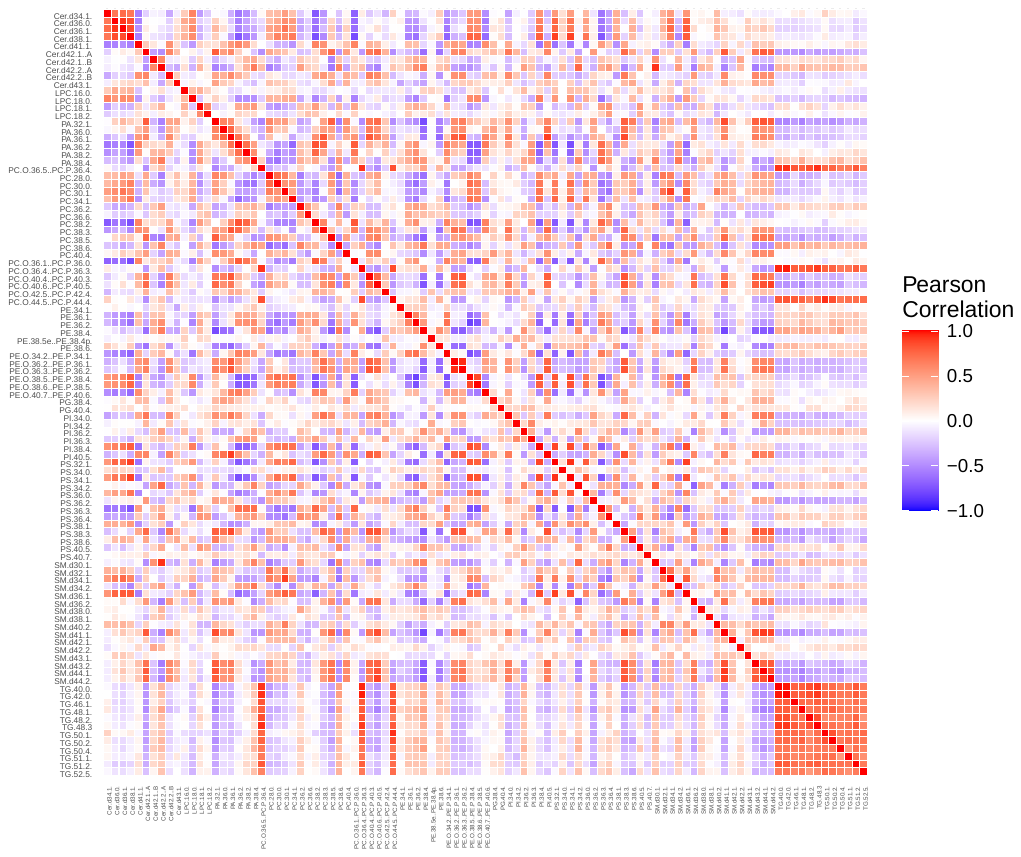

**Supplementary Figure 1. Heatmap depicting the pairwise correlations between lipids.**

CER= Ceramides; SM=Sphingomyelins; PA=Phosphatidic acids, PC=Phosphatidyl-cholines, PI=Phosphatidyl-inositols; PS= Phosphatidyl-serines; PE=Phosphatidyl-ethanolamines; TG=Triglycerides.

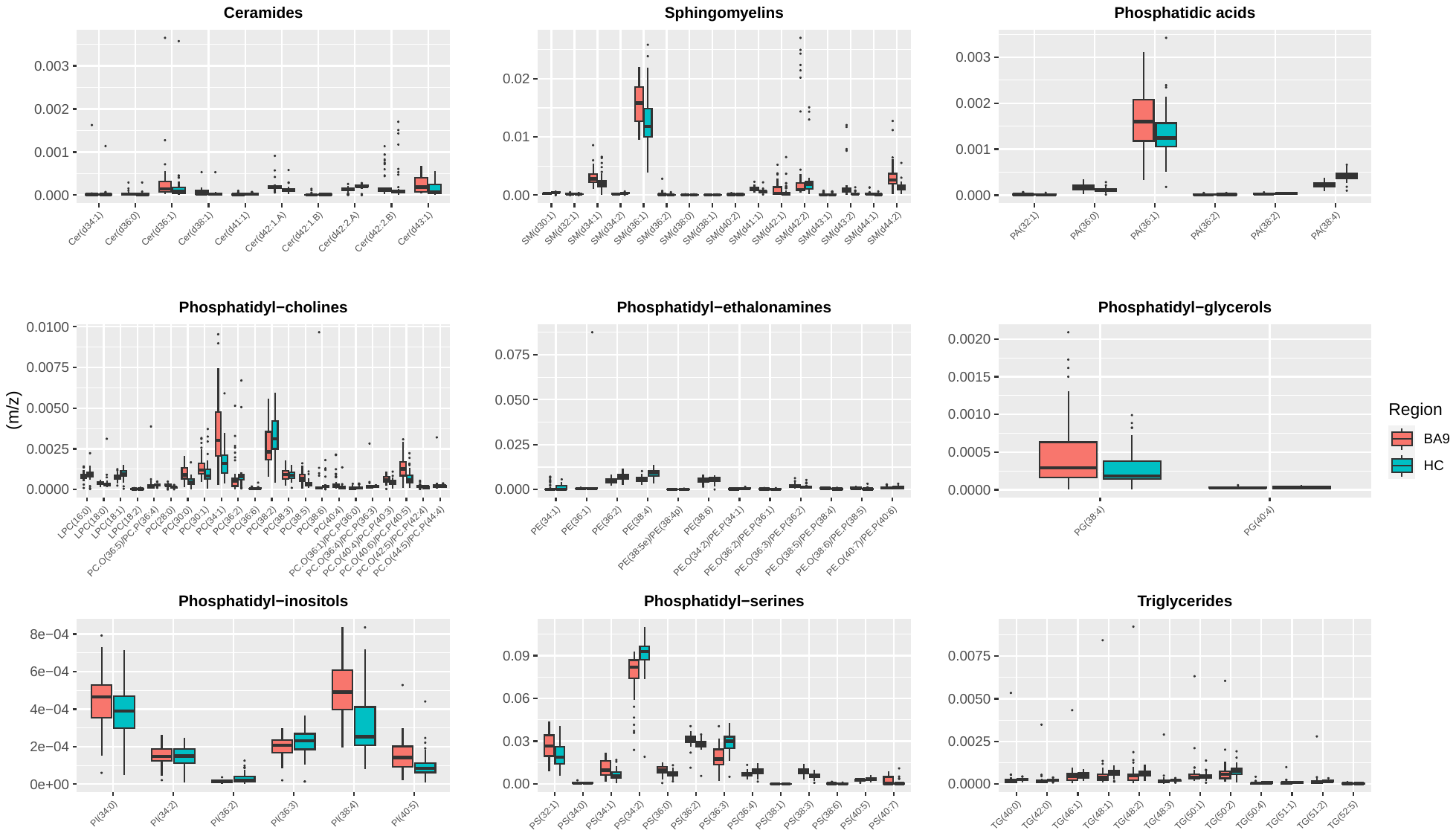

**Supplementary Figure 2. Boxplots depicting raw lipid levels prior to QC in each brain region (BA9 and Hippocampus) separately.**

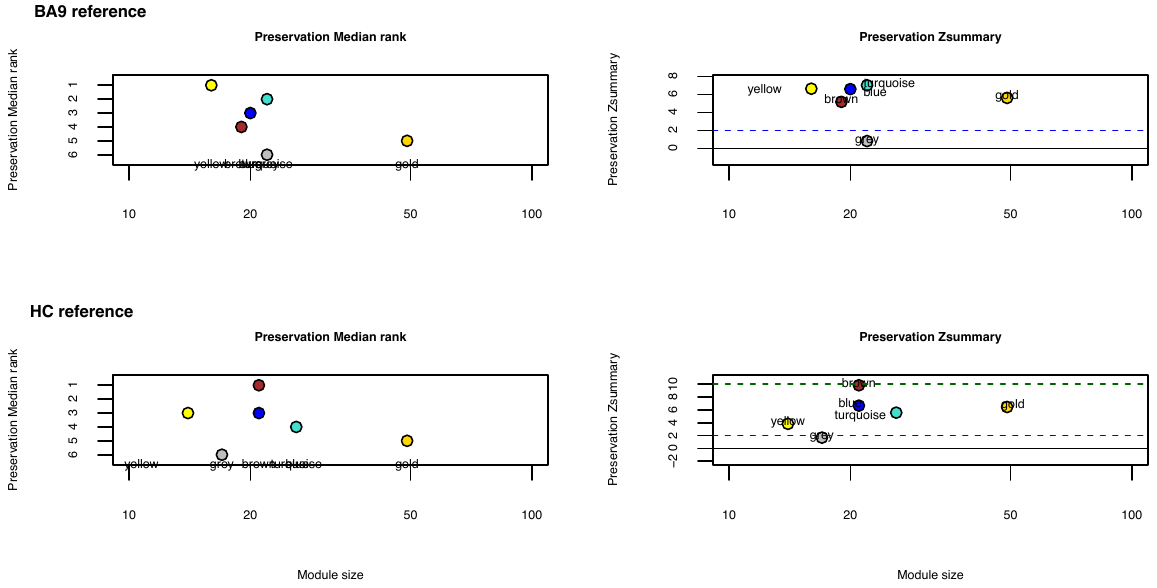

**Supplementary Figure 3. Module preservation between BA9 and Hippocampus.**

Module preservation statistics using a) BA9 as reference and b) Hippocampus as reference.
